# Clinical Feasibility and Preliminary Outcomes of a Novel Mixed Reality Based System to Manage Phantom Pain for Patients with Lower Limb Amputation.: A Pilot Study

**DOI:** 10.1101/2020.08.29.20133009

**Authors:** M. Annaswamy, Kanchan Bahirat, Gargi Raval, Yu Yen Chung, Balakrishnan Prabhakaran

## Abstract

**Objective:** This report describes the details of Mr. MAPP, a virtual mirror therapy system and the results of a clinical feasibility and evaluation study of a pilot sample of patients with history of lower limb amputation and phantom limb pain.

**Materials and Methods:** Mr. MAPP system uses a Kinect V2 RGB-D camera to capture the patient, generates a live interactive 3D model, and incorporates it into a virtual exergame. Mr. MAPP mirrors their preserved limb and allows participants to visualize their amputated limb virtually and perform their prescribed exercise program. Mr. MAPP creates a digital diary of their therapy sessions by recording game performance and errors. Ten consecutive patients with history of limb loss and phantom pain were targeted for this pilot study. They received a Mr. MAPP system and a laptop for home use for 1-month. Outcome measures including pain intensity and pain interference measured on Numerical Rating Scale (NRS) and McGill Pain Questionnaire (MPQ) as well as functional outcomes measured on Patient Specific Functional Scale (PSFS) were obtained at baseline, and after 1-month.

**Results:** Seven participants were enrolled, out of which four completed the study. The study was temporarily suspended due to restrictions imposed during the COVID-19 public health emergency. In-game performance data showed a trend towards improvement in most participants. Compared to baseline, NRS and MPQ scores at 1-month showed no clear trends. PSFS scores improved for 1 participant and remained unchanged for others.

**Conclusion:** Gamified home exercises performed via a novel, mixed-reality system—Mr. MAPP— using which virtual mirror therapy was delivered, is feasible and has potential to improve pain and pain related function in patients with lower limb amputations and phantom limb pain.

## Introduction

Following a limb amputation, the amputee commonly experiences a phantom sensation of a still intact amputated limb. Phantom limb pain (PLP) is defined as a painful phantom sensation perceived in the amputated part of the body.[1, 2] PLP is frequently chronic (>3 months in duration) and is considered one of the most traumatic consequences of amputation, reported in 50%–85% of amputee patients.[3, 4] Patients with PLP have qualitatively described it as throbbing, “pins and needles”, shooting, stabbing, burning, or telescoping, gnawing and cramping.[5-7] The type, frequency, and stability of perception can vary.[8] The diverse nature of PLP makes its treatment challenging.

More than 25 treatments for PLP are currently available, yet there is no consensus choice or a clearly superior option.[9-11] Several approaches have been recommended: behavioral (mirror visual feedback, motor imagery, prosthesis use), psychological (cognitive behavioral therapy, hypnosis, eye movement), surgical, minimally invasive, pharmacological and psychophysical. A review of efficacy of treatments including targeted muscle reinnervation, repetitive transcranial magnetic stimulation, imaginal phantom limb exercises, mirror therapy, virtual and augmented reality (VR and AR), and eye movement desensitization and reprocessing therapy revealed that none of the above treatments is consistently better than a control condition.[12] No treatment has proven to be effective long-term either.[13-15] In order to help maintain effectiveness long-term, better adherence of patients in consistently performing their home exercise program (HEP) seems essential to achieve sustained central nervous system modulation.[16] There is little high quality evidence of efficacy for PLP treatments and most treatment trials are underpowered due to small sample sizes. The lack of efficacy likely speaks to the multiple mechanisms that contribute to PLP both between and within individuals who have sustained an amputation.

A classic non-pharmacological approach for treating PLP is mirror therapy (MT).[17] MT provides this feedback by arranging a mirror (sometimes in a box, known as mirror box therapy [MBT]) in such a way as to allow amputees to view a reflection of their anatomical limb in the visual space occupied by their amputated or phantom limb.[18, 19] MBT utilizes a cardboard box which is open from the top. A mirror is placed at the center of the box. The patient is asked to place their intact limb in front of the mirror and affected limb behind it such that an image of the intact limb is perceived as the amputated limb. The movement of the unaffected limb during MT induces vivid sensations of movement from the missing muscles and joints of the amputated limb, thereby reducing phantom limb sensation and PLP in some patients.[17, 18, 20, 21] MT and MBT is however constrained by limited spatial movement, requiring a patient to remain in a relatively still posture with the head oriented towards the mirror and the body held in mid-sagittal plane with the mirror.[22] It also requires that the user attempts to ignore the intact limb providing the reflection, in order to focus on the reflected image. Furthermore, the illusion obtained is often wavering. These barriers may result in tedious and stressful therapy sessions, which may substantially reduce treatment compliance.

To overcome these challenges, various virtual and mixed reality-based frameworks based on recent advances in technology have been proposed as potential solutions.[22, 23] O’Neil et al. proposed a virtual MBT which uses a pre-defined 3D model of the arm.[24] Animation of the virtual arm is controlled by a wireless glove put on the patient’s intact limb. Sensors embedded in the glove enables capture of the intact limb movement and its application to the virtual phantom limb. Desmond et al. demonstrated a potential use of this virtual MBT to treat phantom pain.[25] Instead of capturing the intact limb movement, Cole et al. captured the movement from the remaining portion of the affected limb using motion capture devices.[26, 27] However, due to sensing limitation of the motion capture device, only coarse movements could be detected; finer movements were pre-animated, which affected the voluntarily movement. The above two approaches rely on a flat screen for rendering the phantom limb illusion. On the other hand, Murray and colleagues designed a system where the virtual environment is rendered on the head-mounted display. Motion of the intact limb is captured using body sensors attached to different joint of the intact limb which is overlaid onto a pre-built 3D model of the phantom limb. A myoelectrically controlled AR environment (MCARE) was proposed by Ortiz-Catalan et al. which captured the patient and surrounding environment with the help of a standard webcam.[28, 29] Pre-built 3D models of the phantom limb was augmented with the real-time capture from webcam and visualized on a flat screen. The 3D model of the phantom limb was animated using motion information captured with eight bipolar electrodes and markers placed on the stump. In et al. proposed a virtual reality-based reflection system that captured the intact lower limb with a camera from the top.[30] The image of the intact limb was then projected at the corresponding position of the missing limb on the screen creating a symmetrical illusion. However, any mismatch in the movement of the affected limb could potentially disturb the illusion. Thus, it limited free and natural movement. Also, it did not provide any interaction with a virtual object.

The above methods utilized pre-built 3D models of the phantom limb which can notably reduce the sense of embodiment due to misalignment with the physical body part, artificial and rigid look, and mismatch with skin tone and clothing. Moreover, these methods require patients to wear body sensors, electrodes or wireless gloves for capturing movement information. It can potentially restrict voluntarily movement, and cause discomfort or skin irritation. To overcome these challenges, Bahirat et al. proposed a <u>Mi</u>xed <u>Re</u>ality based framework for <u>MA</u>naging <u>Ph</u>antom <u>Pa</u>in (Mr. MAPP).[31] It generates a 3D model of the phantom limb in real-time by capturing the 3D data corresponding to the intact limb with off-the shelf RGB-D cameras and mirroring it. It also enables phantom limb interactions with virtual objects providing realistic and engaging experience. The detailed summary of these methods and their respective characteristics have been previously published.[32]

The primary goals of this clinical pilot study were to evaluate the feasibility and preliminary outcomes of a home-based virtual mirror therapy program delivered with the Mr. MAPP system in a sample of patients with lower limb amputation. We plan to use the results from this clinical pilot study as preliminary data to inform regarding a fully powered prospective, controlled, clinical trial to determine the effectiveness of a home-based virtual mirror therapy program Mr. MAPP in treating patients with limb loss suffering from PLP.

## Materials and Methods

### Mr. MAPP System Overview

The Mr. MAPP framework is designed to create a visual cue by generating corresponding phantom limb in real-time. It leverages widely available RGB-D cameras such as Microsoft’s Kinect (Figure 1).[32] It utilizes the RGB, Depth and Skeleton streams obtained from Kinect V2. Specifically, the raw RGB and Depth streams are integrated to create a textured 3D mesh corresponding to the intact limb.[33] Making use of the symmetry in human body about a medial axis, the skeleton stream provided by Kinect SDK is used as an axis of symmetry for mirroring the intact limb. The 3D mesh of the intact limb is then mirrored about a medial axis to create an illusion of the phantom limb. Moreover, to facilitate the interaction of phantom limb with virtual objects, the skeleton part corresponding to the intact limb is also mirrored.

**Figure 1:**
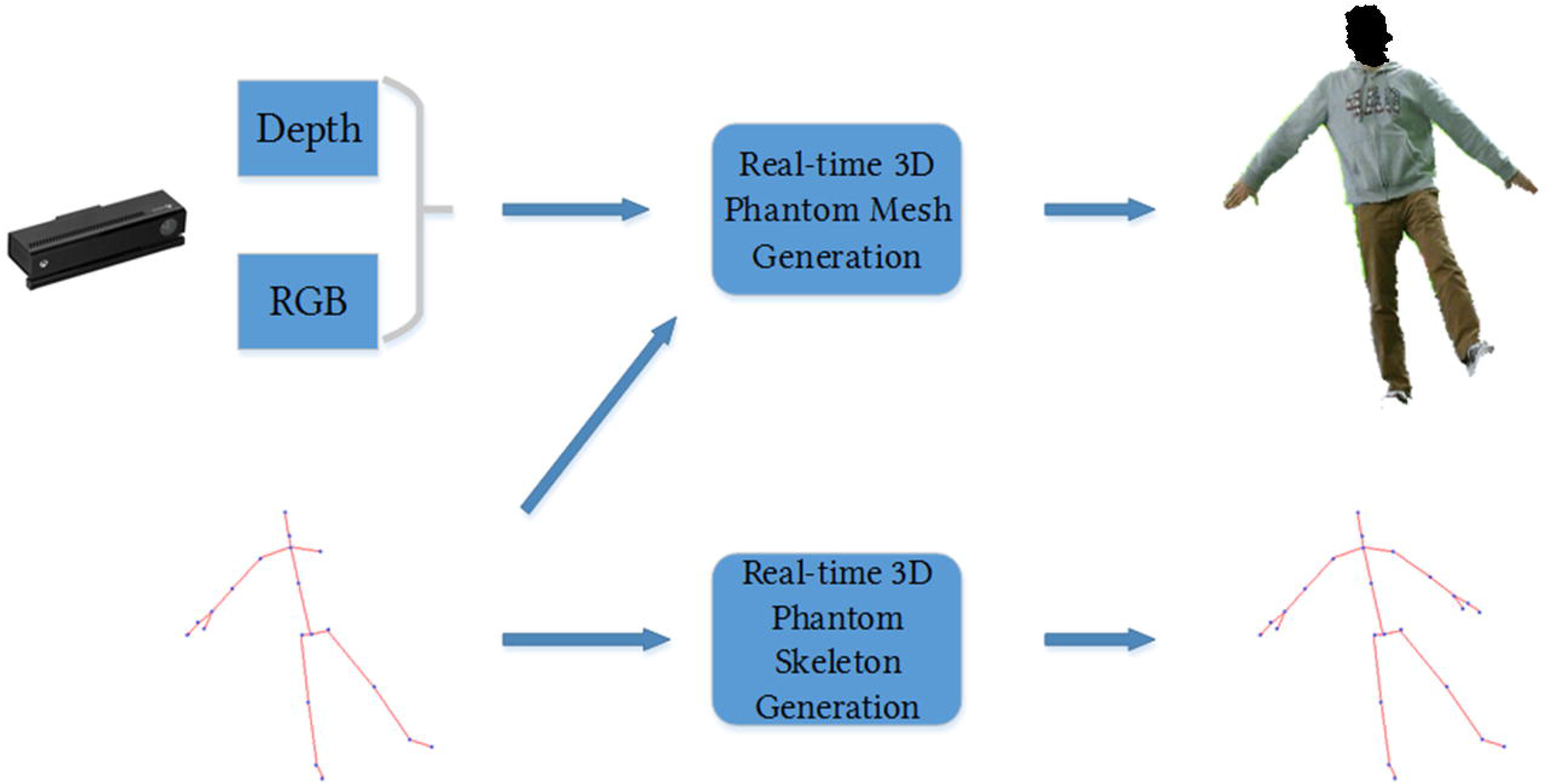
Pipeline of the Mr.MAPP Framework[32]

#### Real time phantom mesh generation

Figure 2 shows the block diagram for the real-time phantom mesh generation outlined in Mr, MAPP framework. It uses the body skeletal joints information provided by Kinect SDK to identify the depth image corresponding to the person. This filtered depth data and camera intrinsic parameters are used to back project each depth point into a real world to obtain a 3D point cloud corresponding to the person. Further, to identify the points belonging to the intact limb and affected limb, the correspondence between 3D points and skeletal segments is established by performing the skeletal segmentation of the scanned point cloud as described in these reports.[34, 35] Points corresponding to the affected limb can possibly interfere with the mirrored data and therefore need to be removed apriori. Further, a 3D representation of phantom limb is crafted by mirroring the 3D mesh of the intact limb about the medial axis, followed by post processing step of hole filling. Detailed description of each step can be found in this paper (Figure 2).[31]

**Figure 2:**
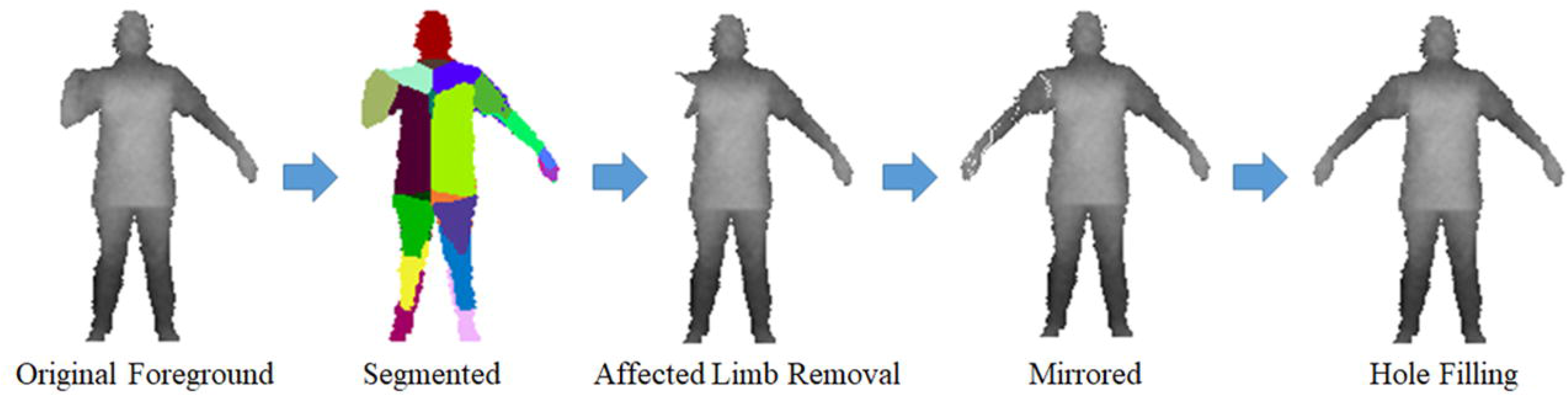
Block diagram for phantom limb mesh generation[31]

#### Extension for lower limb amputation

Even though most of the steps in Mr. MAPP framework can be easily deployed for different limb amputations, a few minor adjustments are required for using it to generate phantom lower limb illusion (Figure 3).[32]

**Figure 3:**
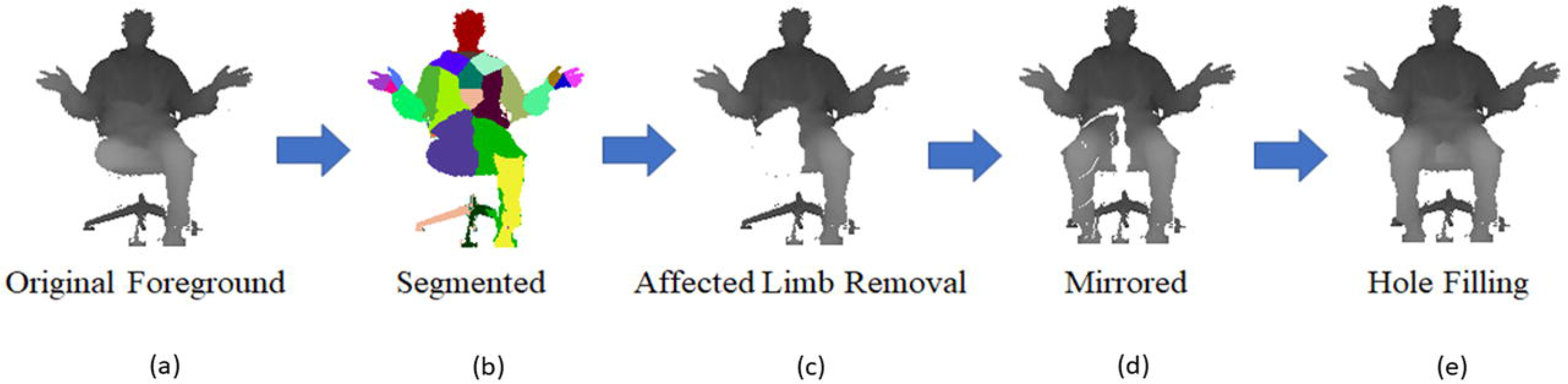
Block diagram of real-time lower phantom limb generation[32]

In summary, the following three major issues are addressed to create realistic phantom limb illusion in case of lower limb amputation with the Mr. MAPP framework:

1. <u>Redefine skeletal coordinate system:</u> If the skeletal coordinate system is defined at ‘spine’ joint, any upper body movement will result in unnatural movement of the lower phantom limb. Hence, skeletal coordinate system must be redefined with ‘hip’ joint at origin.
2. <u>Jittery skeleton:</u> In sitting posture, the skeleton estimation is jittery. Such instability is handled by smoothing skeletal joints using the approach described in [36].
3. <u>Asymmetric segmentation:</u> Inaccurate skeletal estimation leads to asymmetric left and right lower limb region. Such a misalignment between affected and intact limb (Figure 3b) may create gaps when the affected limb is replaced with the mirrored data (Figure 3d). Hence, the root of the affected limb is identified and fills the whole in the surrounding region is filled with the symmetric region from the intact limb.

### Clinical Pilot Study

#### System Setup and Requirements

Mr. MAPP utilizes Microsoft’s Kinect V2 to capture 3D avatar of the person, a computer for running the games and an Oculus Rift for visualizing the virtual environment. A computer used must have Windows 8 or newer, 64-bit (x64) dual-core processor, 2GB RAM and a USB 3.0 port. Whereas, for the Oculus Rift to work, we need Intel i3-6100 / AMD Ryzen 3 1200,FX4350 or greater processor, NVIDIA GTX 960 4GB / AMD RadeonR9 290 or greater graphics card, 8+ GB RAM, Windows 10 or newer, 1x USB 3.0 port, plus 2x USB 2.0 ports and a compatible HDMI 1.3 video output.[32]

#### Physical Setup

While setting up the system, it is important to make sure that there is enough empty space available to move around (approximately 3 meters x 5 meters). User can set up the safe boundary area using the ‘Guardian System’ provided by Oculus. This initial space configuration will be performed only once at the beginning of the study. Typically, the Kinect is placed at height of 1 meter from the ground and person is asked to sit in the chair approximately at distance of 2.5-3 meters in front of the Kinect. After launching the game and once the user is ready, they need to press a virtual ‘START’ button to begin playing the game. Interaction with the virtual ‘START’ button is achieved via button ‘A’ or ‘X’ on the Oculus touch controller or using the mouse. The interface also provides the ability to teleport user to the center of the virtual environment even if the user is not at the center of the action space at the beginning. Also, the user can refine their position physically by slightly moving the chair. However, it is recommended not to make any larger movements as a safety measurement. During each game, user needs to perform corresponding exercise motion correctly in order to successfully complete the game task.

#### Setting, Participants and Study Criteria

The study was approved by our Institutional Review Board. <u>Inclusion criteria:</u> A total of 10 veterans, men and women, over the age of 18, with lower limb amputations (greater than 3 months post-surgery) with phantom limb pain were recruited for the clinical pilot and feasibility study. <u>Exclusion criteria:</u> Patients with open wounds or active infection in residual or contralateral limbs; history of seizures; visual or cognitive impairment that interferes with ability to interact with, participate in a computerized exercise program; any active cardiac condition or an active medical issue that poses a contraindication to exercise therapy; lives more than 60 miles away from the Dallas VA Medical Center; and history of motion sickness induced by head mounted displays (HMDs) or immersive environment. Any patient experiencing the motion sickness induced by HMDs during the therapy session could opt out of the study.

A Health Insurance Portability and Accountability Act (HIPAA) waiver was obtained using which potential study participants with lower limb amputation who were established patients in our Physical Medicine & Rehabilitation (PM&R) clinics were screened for inclusion in the study. Eligible study participants who met criteria were identified and underwent a basic clinical evaluation by rehabilitation physician to confirm that they met criteria. After the evaluation, the patient provided their informed consent and enrolled into the study.

#### Study Protocol and Intervention

Once consent was obtained, study participants were instructed on safely performing the prescribed exercises using the Mr. MAPP system, independently at home. They initially completed the outcome instrument questionnaires. After completing the questionnaire, they donned the goggles and acquainted themselves with Mr. MAPP by moving around in the virtual world. They were then instructed in their exercise therapy session.

Each exercise therapy session consisted of three sub-sections where the participant played three different virtual games. These virtual games were designed specifically for persons with lower limb amputation. Each game targeted certain types of movement. We focused mainly on three movements: 1) knee flexion and extension, 2) ankle dorsiflexion and plantar flexion, and 3) tandem coordinated bilateral lower extremity movement. Each game required about 5 minutes to complete. Participants were instructed to play 2 sessions per day for 1-month.

The research team then provided the Mr. MAPP system including a laptop, and camera and instructed the patient in its use at their home. The participant then used the system to perform their personalized home exercises daily for a 1-month period (to evaluate sustainability of exercise behavior). At the end of this period, the system was returned.

#### Outcome Measures

At initial clinic visit, 1-week and 1-month visit, study participants completed questionnaires of outcome instruments. Primary outcome measures were Pain score on 0-10 Numerical Rating Scale (NRS), McGill Pain Questionnaire (MPQ) and Patient Specific Functional Scale (PSFS).

The participants rated their current average pain on a 0-10 NRS at each assessment.

Study participants provided a more detailed assessment of their pain using the MPQ at each assessment. The MPQ is a classic pain assessment questionnaire that can be used to monitor the pain over time and to determine the effectiveness of any intervention.[37] It was initially developed by Dr. Melzack at McGill University in Montreal, Canada. The three sections of the MPQ are: (1) What Does Your Pain Feel Like? (2) How Does Your Pain Change with Time? and (3) How Strong is Your Pain?

The PSFS is a useful questionnaire that can be used to quantify activity limitation and measure functional outcome for patients with any physically disabling condition.[38] At initial assessment, the participant is asked to identify up to three important activities that they are unable to do or are having difficulty with as a result of their limb amputation and PLP. For each identified activity, they rated their level of functional difficulty on a 0-5 scale. At each follow-up assessment, participants, re-rated their level of functional difficulty on the same 0-5 scale for each identified activity. The total score = sum of the activity scores/number of activities. The minimum detectable change (90% CI) for average score is 2 points, and minimum detectable change (90% CI) for single activity score is 3 points.

Participants received weekly telephone support and online ad-hoc technical support as needed. Adverse event data were also collected, and clinicians were informed for any clinical intervention as needed during this pilot study.

Clinical study data were collected and managed using REDCap (Research Electronic Data Capture) electronic data capture tools hosted at the local VAMC. REDCap is a secure, web-based software platform designed to support data capture for research studies, providing 1) an intuitive interface for validated data capture; 2) audit trails for tracking data manipulation and export procedures; 3) automated export procedures for seamless data downloads to common statistical packages; and 4) procedures for data integration and interoperability with external sources.

#### In-Game Data

The following data were automatically recorded into a ‘digital diary’ as users played their prescribed exercise games at home: session time, duration, and gaming score. Through session time and duration as attendance record, each participant’s engagement could be assessed. Also, the attendance record could be used to verify the self-reported exercise diary by the patient. Each participant’s progress in game performance could be evaluating their game score changes over time. During the 1-month period when study participants used the Mr. MAPP system, each participant were instructed to play two sessions every day where each session includes playing three different games: (1) Ankle dorsiflexion/plantar flexion, (2) Knee flexion and extension and (3) Tandem coordinated movement.

Scores obtained for any game over the course of the study can be a good indicator of complexity of the game and comfort with respect to each individual participant. Using this information, we can further fine tune the complexity of each game to make it more engaging and fun for each participant based on their performance history. Therefore, we analyzed the scores obtained by all participants for each game during the study.

## Results

Ten participants were targeted in this clinical pilot study. However, due to restrictions imposed on all clinical research activities at the onset of the COVID-19 public health emergency, the study recruitment has been temporarily halted at 7 study participants. Out of the 7 patients who consented to participate, 4 patients completed the entire study; 3 patients withdrew from the study (one patient did not follow-up; one patient changed his mind; one patient had prosthesis associated pain and decided to withdraw from the study until that issue could be remedied).

Study participants were mostly male 86% (6/7), mean age was 64, and 57% (4/7) were Black. Most of them had transtibial amputations (Below the Knee-57%; 4/7) that were nearly 1400 days old on an average (Table 1).

**Table 1:** Demographics, Amputation History and Study Participation Status

| Participant | Age Range | Sex (Male/Female) | Race/Ethnicity | Type of Amputation | Time since Amputation (Days) | Study Status |
| --- | --- | --- | --- | --- | --- | --- |
| 1 | 56-60 | Male | White | Left Below Knee | 1241 | Failed to Follow-Up |
| 2 | 50-55 | Male | White | Left Below Knee | 4678 | Complete |
| 3 | 70-75 | Male | White | Right Below Knee | 434 | Participant Declined |
| 4 | 70-75 | Male | Black | Right Above Knee | 308 | Completed |
| 5 | 60-65 | Male | Black | Left Above Knee | 1297 | Completed |
| 6 | 60-65 | Male | Black | Left Below Knee | 1410 | Completed |
| 7 | 60-65 | Female | Black | Left Above Knee | 427 | Pain due to Prosthetic |

### In-game Data

The following attributes of participants’ participation in the virtual exercise therapy sessions were analyzed: attendance of therapy sessions, performance improvement over time and effect of session duration on the performance.

The gaming time for participant #4 and #6 are illustrated in Figure 4. Both participants meticulously performed their exercise therapy sessions daily for 1-month. In Figure 4a, participant #4 tended to start the exercise therapy session at the same time every day, but participant #6 varied their start time. In contrast, digital diaries from participants #2 and #5 revealed more sporadic participation in their exercise therapy sessions.

**Figure 4:**
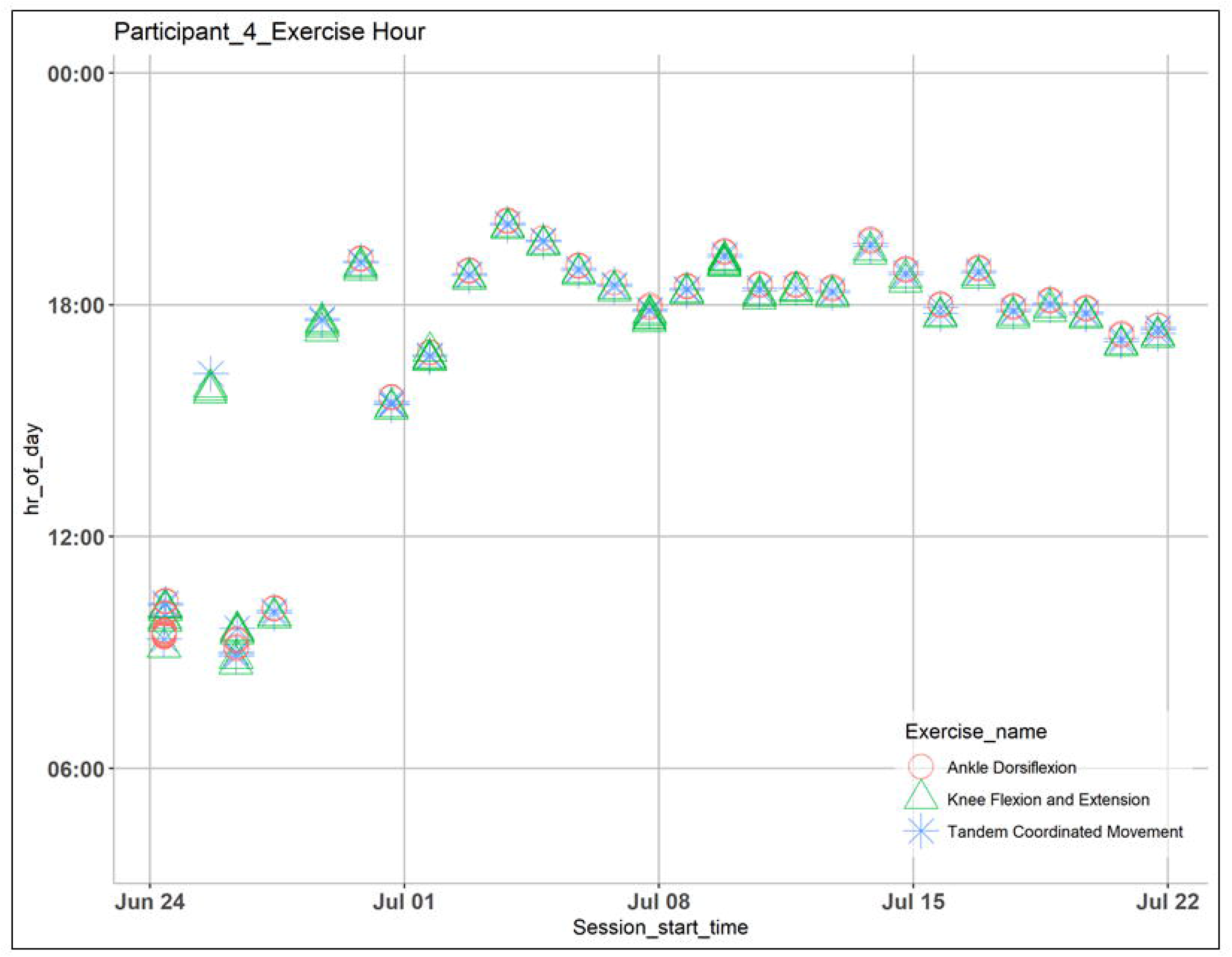
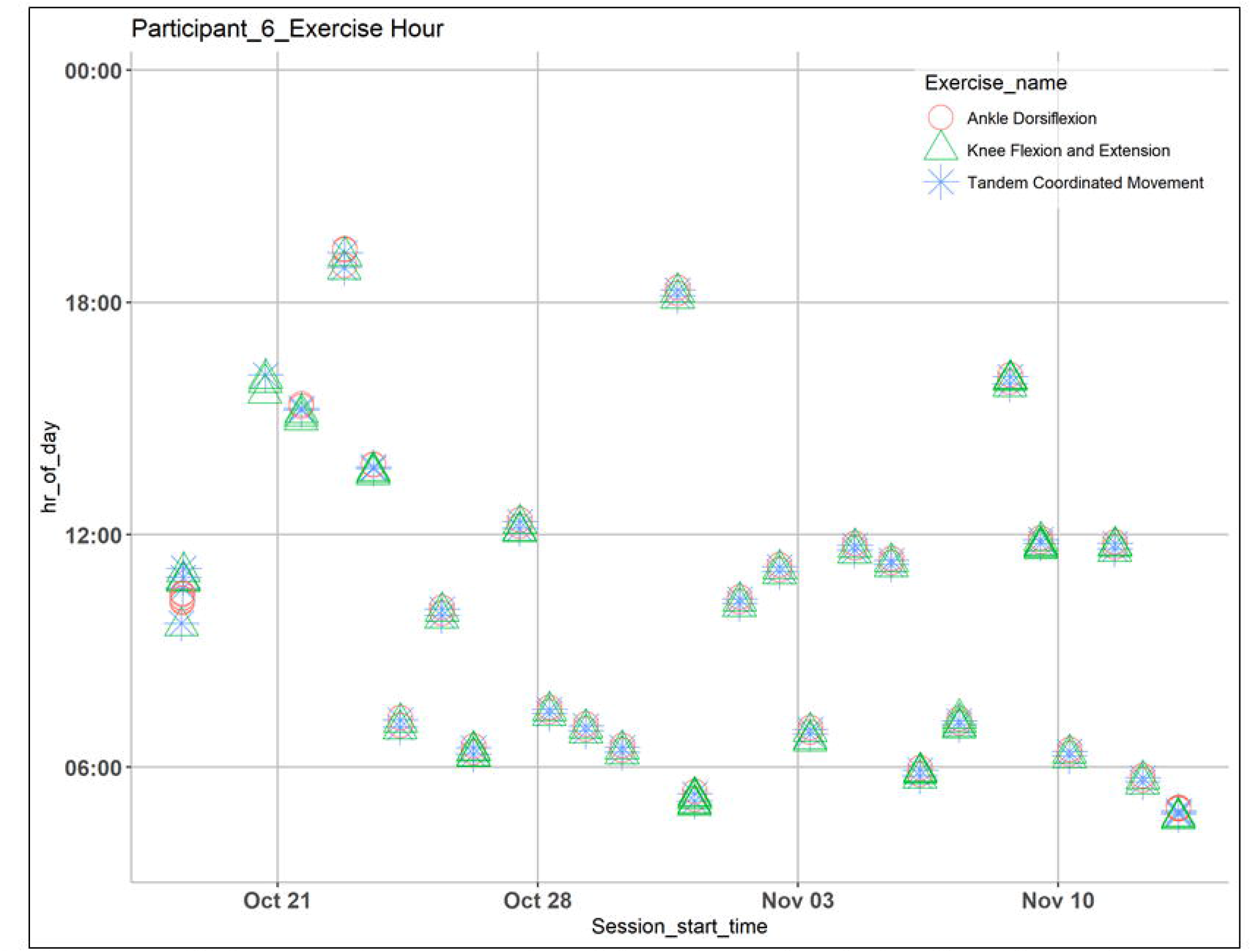
Analysis of time at which a certain game is played for a given day during the pilot study 4a) Participant #4; 4b) Participant #6

The game performance changes over time for each game were evaluated (Figure 5). Each participant’s game performance changes were variable; hence a regression line estimated based on all records across all participants for the game was drawn illustrating a general trend towards improvement across all participants (Figure 5). For example, in the Tandem coordinated movement game (Figure 5a), a positive correlation trend could be observed in participant #4 but the trend was not observed in participant #6. In the Ankle dorsiflexion game, a slight positive correlation was noted in all participants (Figure 5b). For the Knee flexion and extension game the trend is neutral over time.

**Figure 5:**
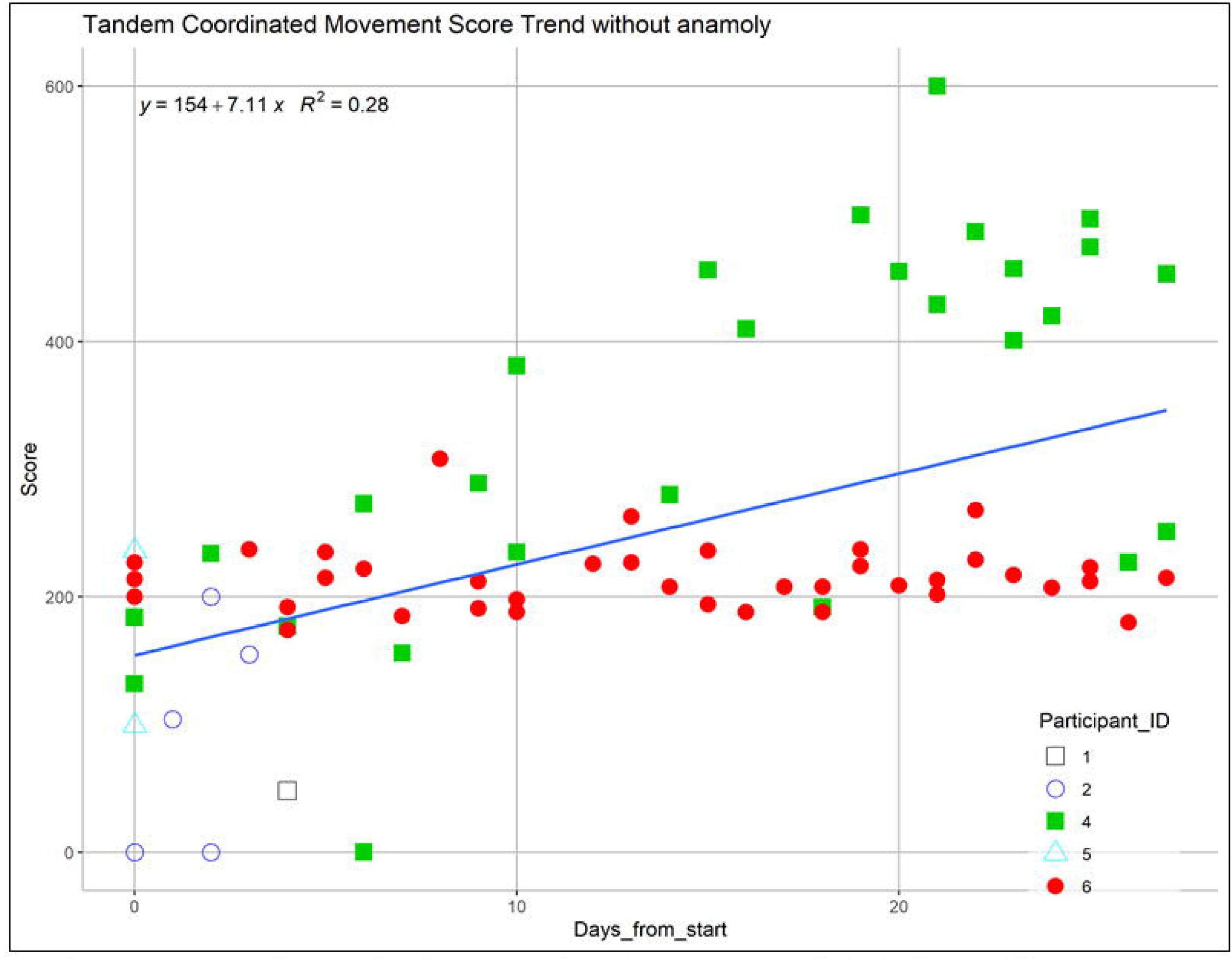
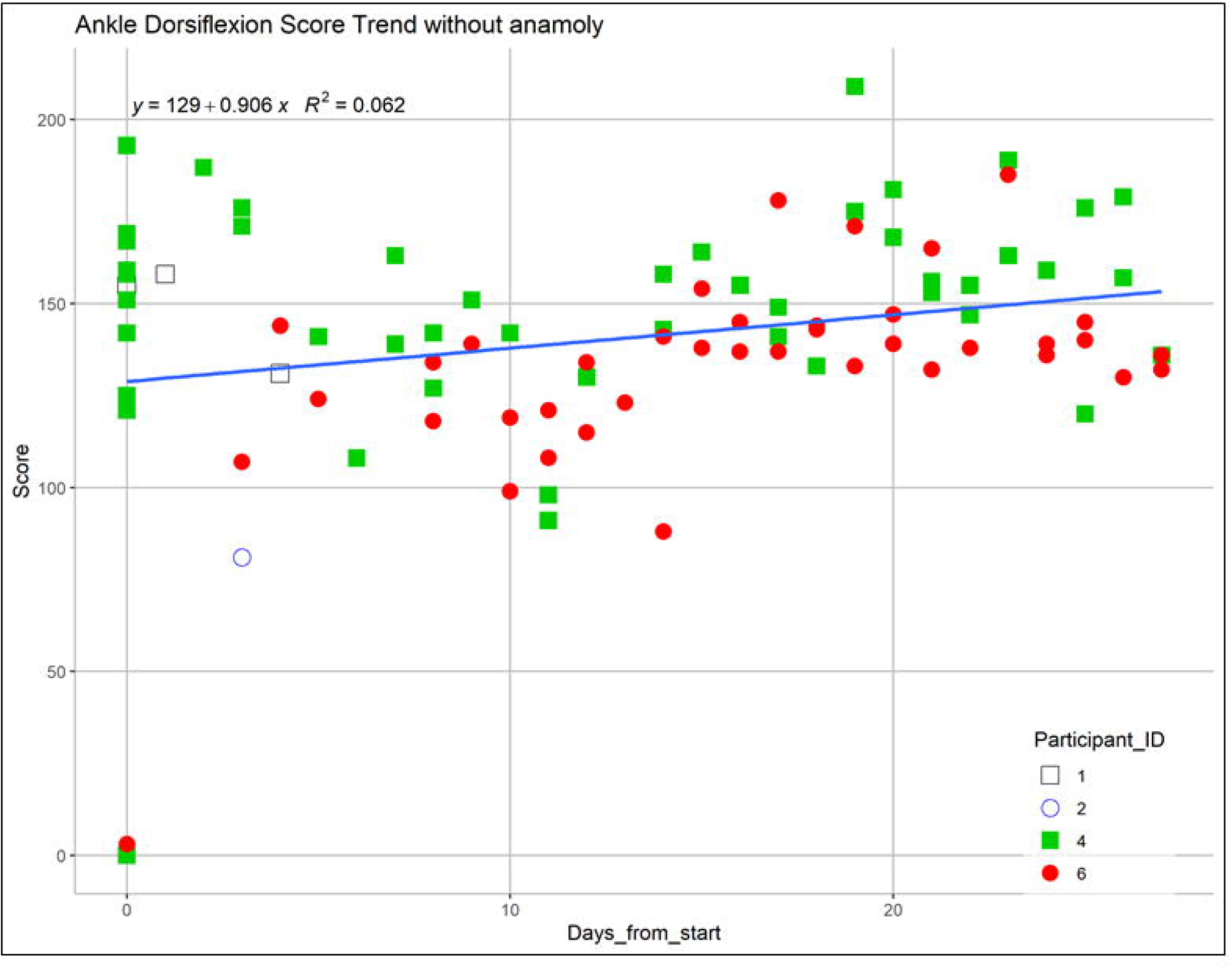
Analysis of scores over the study duration. (Participant #4 is labeled in solid green square, and Participant #6 is labeled in solid red circle) 5a) Tandem coordinated movement; 5b) Ankle dorsiflexion

The relationship between game performance and session duration was also assessed (Figure 6); however, no clear correlation was noted. The Knee flexion and extension game took approximately 2 to 3 minutes to complete but the Ankle dorsiflexion game typically took less than 40 seconds. The Tandem coordinated movement game was not included in this analysis since this game had a fixed duration of 2 mins. However, some data anomalies were noted because with game score was zero in some instances of game play despite several game launches and significant amount of playing time; these data anomalies may have contributed to the lack of correlation.

**Figure 6:**
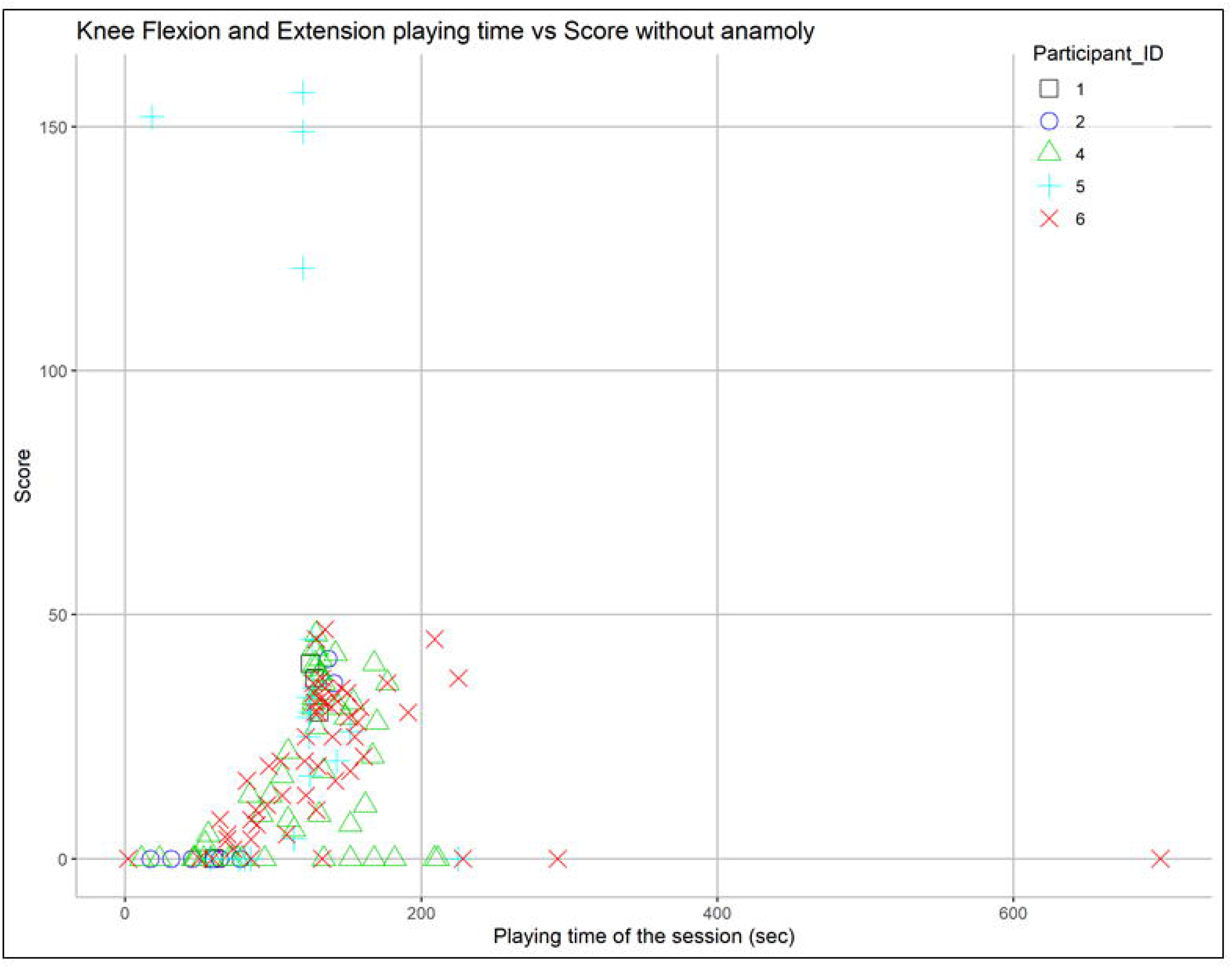
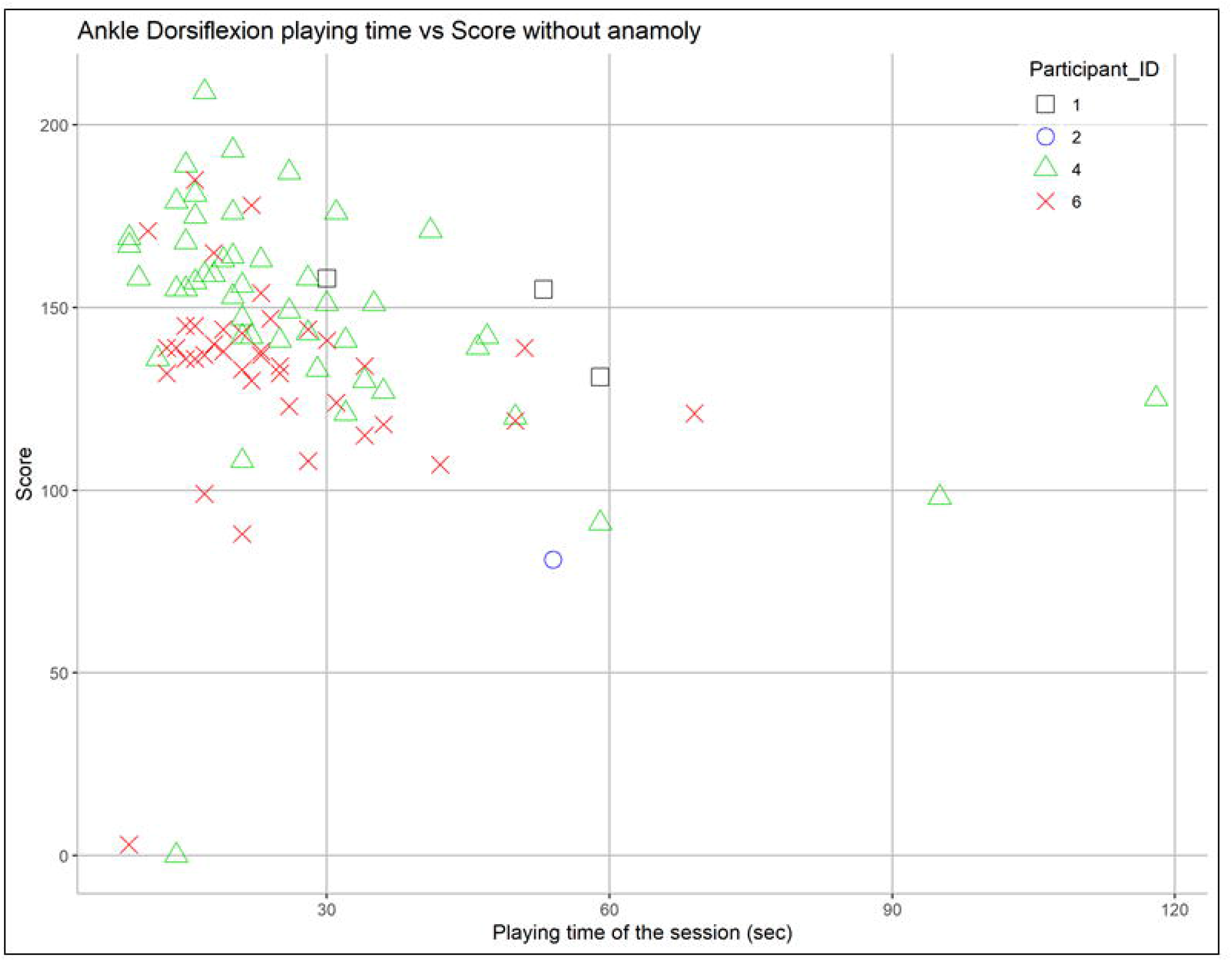
Effect of session duration on the performance 6a) Knee flexion and extension; 6b) Ankle dorsiflexion

### Clinical Outcome Data

Among the 4 study participants who completed the 1-month clinical trial, no clear trends (no improvement or worsening) in change in pain was noted in any of the 3 sections of the MPQ (Table 2) or in pain intensity as rated on the weekly NRS in the exercise diary (paper) (Table 3). However, anecdotally all 4 participants reported that they felt temporary pain relief after playing the exercise therapy games on Mr. MAPP. Participant #2 reported that playing a game during an episode of PLP exacerbation helped to reduce the pain level. No other participant self-initiated such therapy sessions in an attempt to relieve their pain.

**Table 2:** McGill Pain Questionnaire Data

| McGill Pain Questionnaire |  |  |  |  |  |  |  |  |  |
| --- | --- | --- | --- | --- | --- | --- | --- | --- | --- |
|  |  | Section 1: What does your pain feel like? (0-78) | Section 2: How does your pain change with time? | Section 3: How strong is your pain? |  |  |  |  |  |
| Participant | Visit |  |  | Current Pain Strength (1-5) | Pain at its worst (1-5) | Pain at its least (1-5) | Pain of worst toothache (1-5) | Pain of worst headache (1-5) | Pain of worst stomach-ache (1-5) |
| 2 | Baseline | 41 | 2 | 2 | 5 | 1 | 5 | 5 | 3 |
|  | 1-Month | 40 | 2 | 2 | 3 | 1 | 3 | 4 | 4 |
| 4 | Baseline | 45 | 2 | 2 | 5 | 1 |  | 5 | 2 |
|  | 1-Month | 67 | 2 | 1 | 5 | 2 |  | 5 | 2 |
| 5 | Baseline | 40 | 3 | 1 | 2 | 2 | 4 | 4 | 4 |
|  | 1-Month | 38 | 3 | 1 | 2 | 1 | 4 | 4 | 2 |
| 6 | Baseline | 61 | 1 | 2 | 4 | 1 | 5 | 5 | 4 |
|  | 1-Month | 58 | 2 | 1 | 5 | 1 | 5 | 4 | 4 |

Possible responses and key for “How does your pain change with time”:
1 = Continuous and steady
2 = Rhythmic period intermittent
3 = Brief momentary transient

**Table 3:** Exercise Diary Data

| Exercise Diary Data |  |  |  |  |
| --- | --- | --- | --- | --- |
| Participant | Week | Average Minutes Exercised (per week) | Average Minutes Exercised (Total) | Pain Score (0-10) |
| 2 | Week 1 | 25.7 | 18.2 | 4 |
|  | Week 2 | 17.1 |  | 6 |
|  | Week 3 | 15.7 |  | 4 |
|  | Week 4 | 14.3 |  | 4 |
| 4 | Week 1 | 21.6 | 15.4 | 7 |
|  | Week 2 | 12.7 |  | 7 |
|  | Week 3 | 13.7 |  | 6 |
|  | Week 4 | 13.7 |  |  |
| 5 | Week 1 | 47.1 | 42.9 | 0 |
|  | Week 2 | 38.6 |  | 1 |
|  | Week 3 | 40.7 |  | 1 |
|  | Week 4 | 45 |  |  |
| 6 | Week 1 | 40.5 | 28.5 | 3 |
|  | Week 2 | 27.9 |  | 2 |
|  | Week 3 | 23 |  | 2 |
|  | Week 4 | 22.7 |  | 1 |

All 4 participants exercised weekly ranging from an average of 15.4 minutes per week (participant #4) to 42.9 minutes per week (participant #5), with pain scores reported that ranged at the low end between 0-1 for participant #5 and at the high end between 6-7 for participant #4.

PSFS data were available for 3 participants (Table 4). Each participant chose 2-3 activity related goals. Participant #4 and #5 showed no change in PSFS scores (#4: 6.33 to 7.67 and #5: 7.5 to 6.5). However, participant #6 demonstrated significant improvement in PSFS scores: 1 to 3.33; minimum detectable change = 2.

**Table 4:** Patient Specific Functional Scale Data

| Patient Specific Functional Scale (PSFS) |  |  |  |  |  |  |  |
| --- | --- | --- | --- | --- | --- | --- | --- |
|  | Baseline |  |  | 1-Month |  |  |  |
| Participant | Goal 1 | Goal 2 | Goal 3 | Goal 1 | Goal 2 | Goal 3 | Goals: |
| 4 | 7 | 6 | 6 | 8 | 7 | 8 | 1) Be able to stay standing while cooking 2) Ability to clean the house comfortably 3) Walking with crutches with more stability |
| 5 | 6 | 9 | - | 7 | 6 | - | 1) Being able to play around with grandkids 2) Getting in and out of the shower |
| 6 | 1 | 1 | 1 | 3 | 4 | 3 | 1) Walking easier without PLP 2) Standing for long periods 3) Staying asleep without PLP disturbance |
| Range: 0-10<br>0 = Unable to perform activity<br>10 = Able to perform activity at same level as before injury or problem<br>- = No goal set |  |  |  |  |  |  |  |

*Note: Participant #2 did not complete PSFS*

## Discussion

Results of this study showed that Mr. MAPP, an in-home exercise therapy system with its gamified and interactive interface is clinically feasible and can motivate persons with lower limb amputation and phantom limb pain to be more engaged and perform prescribed rehabilitative exercises at home to help improve their pain intensity and pain-related interference with functioning.

Analysis of Mr. MAPP’s in-game digital diary revealed that participants tend to improve their gaming performance mediated mainly through increase in number of repetitions. This suggests that the gamified exercise therapy delivered with Mr. MAPP is challenging and can motivate the participants to improve their exercising behavior over time.

The level of engagement varied from participant to participant. Some participants followed the prescribed exercise regularly, but others did not. A couple of technical factors may have affected engagement. First, positioning of the Kinect camera to accommodate for the participant’s unique home environment constraints may have resulted in a sub-optimal set-up resulting in reduced quality of gaming experience. For example, one participant had a shelf capable of housing the camera only at a height of 1.5m from the ground, which is about 50% higher than the ideal height for camera placement; accordingly, the reconstructed images were a little distorted for this participant. Second, in some cases the available space to exercise was narrow, such as a narrow aisle in their bedroom, which resulted in movement restriction and resultant poor experience.

Other environmental factors affected participant engagement as well. One factor was whether there was any help available to operate the laptop. The game launch interface was on the laptop, but the playing area typically is around 2m distance from the laptop and Kinect camera. Thus, without an assistant who could help in operating the laptop for each session, the participant needed to take off the HMD goggles, operate the laptop, and then move themselves back to the playing area, re-don the HMD goggles, and then play. Some participants played the games while seated in their wheelchair since they needed to move back and forth to operate the laptop when performing prescribed exercises.

Some unexpected issues were noted in the digital diary score record, where some sessions resulted in gaming scores of ‘zero’. Two possible reasons for these issues include: a) orientation of Kinect or Oculus sensor or position of the player was changed. During the initial home setup, the technician would test the displacement of all devices until the game could work. However, after the initial setup if the relative orientations and locations changed, the skeleton collider might not align well with the reconstructed visual limb, resulting in the participant being unable to interact the visible virtual limb with the virtual bubbles, or in the worst case scenario, unable to generate the virtual limb in the virtual environment. b) the teleport procedure might not stable because the target position for teleportation was determined based on the detected skeleton in run time. The goal of teleportation at the beginning of the game is to bring the user to center of the game for interacting with virtual objects. However, the estimated center may vary due to the difference skeleton detected in slightly different postures. Typically, participant could self-correct this issue by moving around slightly to get closer to the gaming center. However, if misplacement is in the vertical direction, this discrepancy cannot be corrected, resulting in the participant not being able to play the game normally. Optimizing the above technical and environmental factors is important in the future, as we plan fully powered clinical trials with Mr. MAPP, so as to maximize patient engagement and quality of experience which will likely lead to better clinical outcomes.

Study participants who completed the in-home trial with Mr. MAPP did not show any clear trends in pain improvement assessed on the MPQ or the NRS. However, anecdotally all 4 participants reported satisfaction with the system and temporary relief of pain after participating in the exercise therapy sessions with Mr. MAPP. One participant also reported benefit by using the system during episodes of phantom pain, which were instances of self-initiated treatment. Due to the intermittent and episodic nature of phantom limb pain in many patients, self-initiated treatments during episodes can be of value to patients and needs to be further studied. No participant showed significant worsening in their pain outcome scores. These data suggest that performing rehabilitative exercise therapy sessions using Mr. MAPP system has potential to improve phantom pain although no sustained pain improvement was noted in this 1-month clinical pilot trial. Self-initiated treatment sessions to alleviate pain during phantom pain episodes may be a feasible option for patients with PLP if this therapy modality was available as needed at their disposal.

Analysis of the exercise diary data suggest that participants who completed the study were able to adhere to their recommended exercise therapy protocol. Mr. MAPP system provided them the motivation and incentive to perform their prescribed exercise therapy, and some were motivated to perform additional sessions as well. Occasional discrepancies were noted between digital diary record and paper exercise diary data, likely because of recall errors or participants including non-game exercises also in their paper exercise diary record.

Analysis of PSFS scores indicated that there was no detectable change in 2 participants, but one participant made significant functional gains with the use of the Mr. MAPP system for 1-month. These data suggest that there is opportunity to improve in physical functioning in patients with phantom pain. The likely mediator of this effect is pain related interference with functioning; if pain intensity is lessened, and pain related interference is alleviated, pain related functioning improves as a result. Additional evaluation of the effect of Mr. MAPP in pain, function and pain-related functioning would help understand the mediators of functional improvement in patients with limb loss and phantom limb pain. Behaviors that sustain motivation to perform their prescribed exercises, especially if this therapy leads to pain and functional improvement, are desirable.

The potential to improve pain and functional outcomes in patients with chronic limb loss and phantom limb pain, by non-pharmacological and non-invasive methods of treatment with that is home-based is a tremendous opportunity in healthcare. Improving the quality of life and function in this patient population without additional utilization of trained healthcare personnel and with minimal additional cost of care (given no specially trained personnel are required to oversee therapy sessions) provides significant value to their healthcare. Furthermore, Mr. MAPP facilitated home exercise therapy can potentially mitigate the increased utilization of rehabilitation services that is often observed in this population.

None of the study participants reported any adverse events with the use of Mr. MAPP or with any of the exercises they performed. All study participants were satisfied with the ease of use of the system. All participants also expressed the desire to retain the system for a longer trial if that were possible.

Future comparative trials between this system and usual standard of care MT or MBT may help more conclusively demonstrate the benefit of this system for patients with limb loss and PLP. We also plan to evaluate the outcomes of patients who self-initiate additional treatment sessions, and desire to retain the system for longer trial duration. Also, we plan to develop a Mr. MAPP therapy system module for patients with upper limb loss as well. Future, fully powered, randomized clinical trial comparing Mr. MAPP to other standard of care pharmacologic or physical therapy interventions is also being planned.

### Limitations

This was a clinical feasibility and pilot evaluation study of a small sample of patients with phantom limb pain, meaning this study was not powered to measure clinical efficacy. While some mild trends in clinical improvement in pain and functional outcomes were observed, caution needs to be exercised to avoid over-interpreting these outcomes. Also, we cannot clearly attribute the cause of the observed outcomes to the intervention because this study was not statistically powered to evaluate treatment effect and did not have a control group. Several factors that may influence outcomes were not controlled in this study including duration of pain, type and characteristics of PLP, etiology of amputation, or level of amputation. In addition, number of minutes exercises was variable within our pilot sample of participants, which could have influenced outcomes. Although study participants did show increased engagement over time and increasing motivation to perform prescribed therapy was an objective of the intervention, motivation was not directly evaluated in this study. Despite the above limitations, this clinical feasibility pilot study did demonstrate that exercises performed using virtual mirror therapy with the Mr. MAPP system is clinically feasible and shows potential in its ability to improve pain, and pain-related physical functioning outcomes for patients with limb loss and phantom limb pain.

## Conclusions

Using Mr. MAPP, a novel, gamified virtual mirror therapy system to perform exercises at home by community-dwelling patients with limb loss is feasible and has potential to improve pain and pain-related functional outcomes. Further research including a fully powered prospective study with appropriate control is needed to better evaluate its efficacy in improving pain and pain related function for patients with limb amputations.

## Data Availability

All data referred to in this manuscript is available and stored in the institutional's data repository.

## Acknowledgements

We thank the contributions of Mitchell Kroll, and Sagnik Dakshit towards this project.

## Notes

### Competing Interest Statement

The authors have declared no competing interest.

### Clinical Trial

ClinicalTrials.gov Identifier: NCT04529083 (Retrospectively registered)
Explanation for why the trial was retrospectively registered: Initial guidance we received was that pilot clinical studies are not considered clinical trials and do not need to be registered as such.

### Funding Statement

No external funding was received for the conduct of this study. One or more authors of this work are/were employed by the US federal government and this work was conducted while they were working for the US federal government.

### Author Declarations

Institutional Review Board of VA North Texas Health Care System approved this study

